# Heritability of cardiovascular health across three generations in South Africa: the Birth to Twenty-Plus cohort

**DOI:** 10.1101/2022.03.18.22272642

**Authors:** Lisa J Ware, Innocent Maposa, Andrea Kolkenbeck-Ruh, Shane Norris, Larske Soepnel, Simone Crouch, Juliana Kagura, Sanushka Naidoo, Wayne Smith, Justine Davies

## Abstract

**Objectives:** Cardiovascular disease is increasing in many low-middle income countries, including those in Africa. To inform strategies for the prevention of cardiovascular disease in South Africa, we sought to determine the broad heritability of phenotypic markers of cardiovascular risk across three generations.

**Design:** A cross-sectional study conducted in a longitudinal family cohort.

**Setting:** Research unit within a tertiary hospital in a historically disadvantaged, large urban township of South Africa.

**Participants:** 195 individuals from 65 biological families with all three generations including third generation children aged 4-10 years were recruited from the longest running intergenerational cohort study in Africa, the Birth to Twenty Plus cohort. All adults (grandparents and parents) were female, while children were male or female.

**Primary and secondary outcome measures:** The primary outcome was heritability of blood pressure (BP, brachial and central pressures). Secondary outcomes were heritability of arterial stiffness (pulse wave velocity), carotid intima media thickness (cIMT), and left ventricular mass indexed to body surface area (LVMI).

**Results:** While no significant intergenerational relationships of BP or arterial stiffness were found, there were significant relationships in LVMI across all three generations (p<0.04), and in cIMT between grandparents and parents (p=0.0166). Heritability estimates were 23-44% for cIMT and 21-39% for LVMI.

**Conclusions:** Structural indicators of vascular health, which are strong markers of future clinical cardiovascular outcomes transmit between generations within African families. Identification of these markers in parents may be useful to trigger assessments of preventable risk factors for cardiovascular disease in offspring.

**Strengths:**

- Intergenerational transmission was evaluated for a range of indicators of cardiovascular health within urban African families
- The sample included biological family members from three generations
- Heritability estimates were compared for three commonly used statistical methods.

**Limitations:**

- The sample size is a limitation with the random family statistical method used to increase the numbers of comparisons available.
- Only maternal family members were included.

## Introduction

Within South Africa, a quarter of all adults are hypertensive and one in five deaths are from cardiovascular disease (CVD)^1^. CVD mortality and morbidity are set to rise with increasing life expectancy (now at 64 years; an increase of 10% in the last decade)^2^, and increasing levels of overweight and obesity (68% women, 31% men)^3^. Much focus is placed on detecting and treating CVD, but with limited healthcare resources, pragmatic approaches are needed including primary prevention in younger, at-risk individuals to prevent CVD^4^.

There is evidence that strong predictors of future adverse cardiovascular outcomes (such as heart attacks and strokes) may be transmitted through biological families so that measures in parents or grandparents may identify children at future risk^5^. Early vascular predictors of CVD outcomes include both structural (e.g. thickening or stiffening of arterial walls, cardiac hypertrophy) and functional changes (e.g. elevated blood pressure)^6-10^. Hypertension is the largest contributor to CVD in Africa, with research showing elevated blood pressure in children as young as 5 years of age^11^. Studies of mono- and dizygotic twins have shown high heritability of systolic and diastolic blood pressure in populations of both African and European decent^12 13^, though heritability may be lower for individuals of African decent^14^. Within South Africa, data is also emerging that blood pressure is heritable across families (parent-child, and sibling pairs)^15^. However, due to the high levels of hypertension in South African adults, hypertension in a family member is unlikely on its own to be a sensitive enough indicator to identify at risk young adults or children for intervention..

As such, additional measures may be needed to identify those family members most at risk and where early intervention may have greater returns. Evidence from outside of Africa has shown that several other markers of cardiovascular disease risk are heritable. For example, central blood pressures may show stronger heritability than the brachial blood pressures typically measured in routine care;^16^ carotid artery structure, function and pathology have been shown as heritable, with diameter and carotid intima media thickness appearing as the most heritable traits;^17-19^ arterial stiffness, as assessed by pulse wave velocity, has also been reported as heritable within family studies; and ^16 20^ findings from echocardiography studies suggest that several cardiac measurement parameters may be heritable within families, including left ventricular (LV) function and structure including LV mass and LV hypertrophy^21-24^. Indeed, the combination of arterial stiffness and central pressure has been suggested as a potential tool to investigate risk in nuclear families^25^.

However, there is limited evidence from African families to indicate which indicators of cardiovascular health are most related and therefore, potentially most useful to indicate intergenerational risk within family units in South Africa. One previous study suggested that echocardiography may be particularly useful to detect intergenerational transmission of changes in cardiac structure and function in South African families (parent-child, sibling-sibling pairs)^26 27^, though how this and other vascular measures are related across children, parents and grandparents in the region is not known. Additionally, the frequent background of undernutrition and burden of infectious diseases may mean that heritability estimates are different in Africa to elsewhere.

Therefore, we sought to investigate how a range of indicators of cardiovascular health (brachial and central pressures, arterial stiffness, carotid intima media thickness and echocardiography findings) were related within three generations (grandparents, parents and children) of African families from urban South Africa to inform further risk identification and potential targeted CVD prevention efforts.

## Methods

### Study population and sample size

Biological families with three generations (grandmother, mother and child [boy or girl age 4-10 years]) were recruited from the largest and longest running birth cohort study in Africa; the Birth to Twenty Plus (Bt20-plus) cohort described in detail previously^28 29^. A database of 162 index children (now the mothers) was drawn from previous Birth to Twenty studies that indicated index children with survival of their biological mother and birth of a biological child. These index children were then contacted by telephone to confirm the presence of their biological mother, and a biological child between the ages of 4 and 9 years, with eligible families invited to take part. Families with participants who were pregnant, experiencing current acute illness, or with any major congenital disorders were excluded. The study design was a cross-sectional in-depth assessment of vascular health at a research unit located in a large hospital in Soweto. Data was collected between August 2019 and March 2020. Previous work in East African families found high heritability of blood pressure (systolic, diastolic and pulse pressure h^2^ 0.37, 0.24, 0.54), though the authors did not assess other vascular measures^30^. Based on these previous reported levels of heritability between two generations and using the methods of Klein et al.^31^, n=65 families (n=195 individuals) at alpha=0.05, would give 82% power to detect an h^2^ of 0.4, and 94% power to detect an h^2^ of 0.5 in blood pressure. With three generations, these estimates may be conservative.

### Ethical considerations

Trained researchers who spoke the participant’s home language explained the study and all participants provided written informed consent prior to taking part in the study. For children, the mother of the child provided written consent, with children age 7 years and above also giving their written assent to take part. The Human Research Ethics Committee (Medical) of the University of the Witwatersrand approved the protocol (Ref: M190263). We used the STROBE cohort checklist when writing our report^32^.

### Patient and Public Involvement

The study design was informed by previous work with two generations from this cohort, where participants expressed a desire to include additional generations in cardiovascular health assessments. However, participants were not involved in the study design, recruitment or conduct of the study. During 2022, a series of workshops are planned with the community to disseminate results and to explore the co-creation of potential community level interventions.

### Measurements

Standard protocols were used for collection of all data, with the same staff repeating all measures or assessments of inter-operator variability conducted as described further in the appendix. Medical history (including antihypertensive medication use) and health behaviours were recorded via self-report. Tobacco use (daily or occasional current use of both smoked and smokeless tobacco products) was assessed using questions from the Global Adult and Tobacco Survey^33^. Alcohol use was evaluated using the World Health Organization Alcohol Use Disorders Identification Test (WHO-AUDIT)^34^, with hazardous or harmful alcohol use assessed as an AUDIT-C score (first three questions – shortened form) of ≥3 and/or a total AUDIT score of ≥ 8.

Trained researchers measured height and weight in triplicate to the nearest 0.1cm and 0.1kg using a portable stadiometer and electronic scale (SECA, Hamburg, Germany). Waist and mid-upper arm circumference (MUAC) were measured in triplicate to the nearest 0.1cm following standard measurement protocols^35 36^.

All measures were taken in the morning following an overnight fast and with no caffeine or tobacco for at least 3 hours prior to measurement. Using the Sphygmocor Excel device (AtCor Medical, Naperville, USA) with appropriate size brachial cuff, brachial blood pressure and resting heart rate were determined, and central arterial pressures (cSBP, cDBP, pulse and mean arterial pressure) were estimated. Three measurements were taken, with the second and third measures averaged for analysis. Ultrasound measures were taken in triplicate with the Mindray DC-70 Ultrasound system (Mindray, Shenzen China). Further detail for these assessments is provided in the appendix.

### Analyses

The primary outcome was heritability of blood pressure (BP, brachial and central pressures). Secondary outcomes were heritability of arterial stiffness (pulse wave velocity), carotid intima media thickness (cIMT), and left ventricular mass indexed to body surface area (LVMI). All exposure effects were adjusted for age, height, weight and sex in the regression models, with heritability estimates adjusted for age.

For adults, body mass index (BMI kg/height m^2^) was categorised as follows: <18.5 underweight; 18.5-24.9 normal weight; 25.0-29.9 overweight; ≥30 obese. Children’s BMI was categorised as underweight, normal, overweight or obese using age- and sex-specific cut-offs from the International Obesity Task Force (IOTF)^37^. Waist to height ratio was calculated for both adults and children, as this has previously been shown as a predictor of health risks of obesity across the lifecourse in all ethnic groups^38^. In adults, prehypertension was defined as 120-139 mmHg systolic or 80-89 mmHg diastolic and not currently taking antihypertensive medication, while hypertension was defined as a blood pressure ≥ 140 mmHg systolic or ≥ 90 mmHg diastolic or currently taking antihypertensive medication. For children, elevated blood pressure was defined using the age, sex, and height adjusted percentiles of the American Academy of Pediatrics Clinical Practice Guideline (2017)^39^.

The Devereux formula was used to calculate LVM^40^ and left ventricular mass index (LVMI) was calculated as a ratio of LVM indexed to body surface area^41^. Left ventricular hypertrophy (LVH) was defined as LVMI>95g/m^2^ for adult women and LVMI >95th percentile for children. Normality of data were checked with visual inspection of histograms and the Shapiro-Wilk test^42^.

Our analyses followed two stages. Stage 1) determining the association between parent-offspring pairs for each of the vascular health traits, and stage 2) estimating heritability for traits that exhibited an association in the parent-offspring pairs. Participant characteristics and the associated vascular health measurements are also described.

### Stage 1. Random family method

In this study we used the random family method as described in detail by Usuzaki et al. (2020) and implemented the analysis based on Heß (2017) randomization inference algorithm^43 44^. We used resampling of the exposure variable to generate the distribution of parental trait effect on offspring, controlling for confounding variables as below. We used the classical model generally used to explore heritability in phenotypic traits:

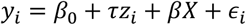

where *y*_*i*_ is the offspring trait, *τ*is the “treatment” effect (regression slope) for *z*_*i*_, the parental trait. *X* is a matrix of control variables and *β*the associated coefficients. *τ*is obtained for the original pairs (*z*_*i*_, *y*_*i*_) and using randomization inference tests, we performed 5000 resampling-based pairs to obtain the distribution of the statistic *τ*, that is, the distribution of random parental trait effect on offspring’s corresponding trait. Randomization inference tests have the advantage that they can handle small sample sizes and do not rely on validity of the specified model regardless of the generated statistic being from the model^43^. Randomization inference also produces the distribution of a test statistic under a designated null hypothesis, thereby allowing us to assess whether the observed (original parent-offspring pair) relationship statistic (regression coefficient) is significantly different and hence the null hypothesis can be rejected in favor of the parental trait having a significant influence on the offspring trait. In brief, regression coefficients were generated for all primary and secondary cardiovascular measures within the biological families: adjusting brachial and central pressures, pulse wave velocity and cIMT for age, height, weight and sex; and adjusting LVMI for age and sex only as it is already indexed to body surface area. Restricted resampling of the data was then employed to generate 5000 random family units ensuring random pairing of parent off-spring biological families. The regression coefficients for each cardiovascular outcome marker were then compared between the family pair and random pairs. Kernel density plots of *τ*-values for original family pairs and random pair *τ*-values were then generated to assess statistical significance of the selected traits.

### Stage 2. Heritability Estimation

For those variables which showed significantly greater association between family members compared with randomly generated pairs using the random family method, heritability estimate(s) were derived using the variance components decomposition method based on the linear mixed effects model (LMM) as all vascular health traits of interest were continuous. The Restricted Maximum Likelihood (ReML) method was used to estimate the variance components and hence heritability. However, due to concerns by Hadfield (2010) and Morrissey (2010) on ReML limitations^45 46^, we additionally implemented the Bayesian method for variance components and heritability estimation^47^, thereby creating a range for each heritability estimate. The basic model (LMM) is:

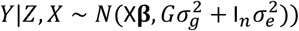

where additive genetic variance of the trait *G* is estimated using relatedness information between individuals or genotypes *Z* with both fixed effects *β* for *X* control variables, 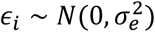 and random effects following a normal distribution with mean 0 and variance 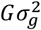^49^. *G* is the genetic relatedness matrix (GRM) and was estimated using the kinship package in R (R version 4.0.2)^49^. We also used the kinship package to plot the pedigree of one family in our dataset. The Bayesian linear mixed model with polygenic effects (*g*) having the following sampling model:

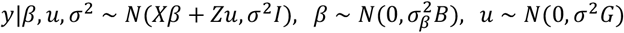

where *B* is known and non-singular diagonal matrix and 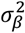 as a hyperparameter was used. The *G* in *σ*^2^*G* is the genetic relatedness matrix estimated through the kinship package for the family relatedness. Note, for this model the likelihood and assumed priors were:

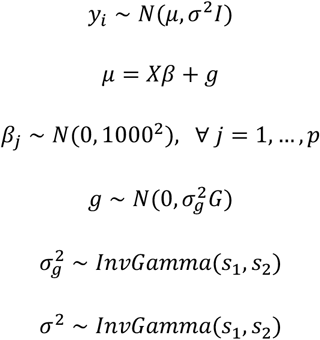

where *s*_1_ *and s*_2_ are chosen to provide noninformative priors. We used rJAGS and rSTAN to perform markov chain monte carlo (MCMC) and hamiltonian monte carlo (HMC) simulations _*σ*_respectively^48^. Heritability was then computed as 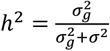. The marginal distributions of all parameters and estimation of the best linear unbiased predictions (BLUP) for the model were obtained using Gibbs’ sampling (MCMC) and the leapfrog integration method (HMC). The samplers made 100000 simulations and only results of the last 90000 were used in the inference. We used two Bayesian paradigms to enable comparisons and manage the inherent uncertainty associated with estimating genetic variance components^45^ as well as in using small sample sizes. Age of the participant was used as a control variable for all models and was standardized together with the vascular health traits before estimation to improve efficiency of Bayesian sampling.

## Results

Of the 162 index children identified: n=48 (30%) could not be contacted either as the telephone number had changed or they did not respond to calls or voice messages; n=14 (9%) did not wish to take part; n=5 (3%) were not eligible due to current illness, pregnancy, or a biological child not in the required age range; n=4 (2%) were no longer residing in Soweto; n=3 (2%) were not available due to school or work commitments; and n=9 (6%) booked appointments but did not attend. Finally 65 families (49% of those contacted) took part in the study providing n=130 adults and n=65 children and generating 195 biological pairings: 130 first generation and 65 second generation).

Whole family completion rates for the vascular measures were as follows: carotid ultrasound (n=63); brachial blood pressure, heart rate and pulse wave analysis (n=62); echocardiography (n=59); PWV (n=40); all vascular measures (n=40). Families with complete anthropometry data and at least one vascular measurement complete for a family pairing (parent/child, grandparent/parent or grandparent/grandchild) were included in the analysis as the random family method does not require all three generations to have data, only that a family has one or more biological pairs with valid measurements. Descriptive characteristics are presented in Table 1, including the number of adults and children with successful measurements for each variable.

**Table 1.**
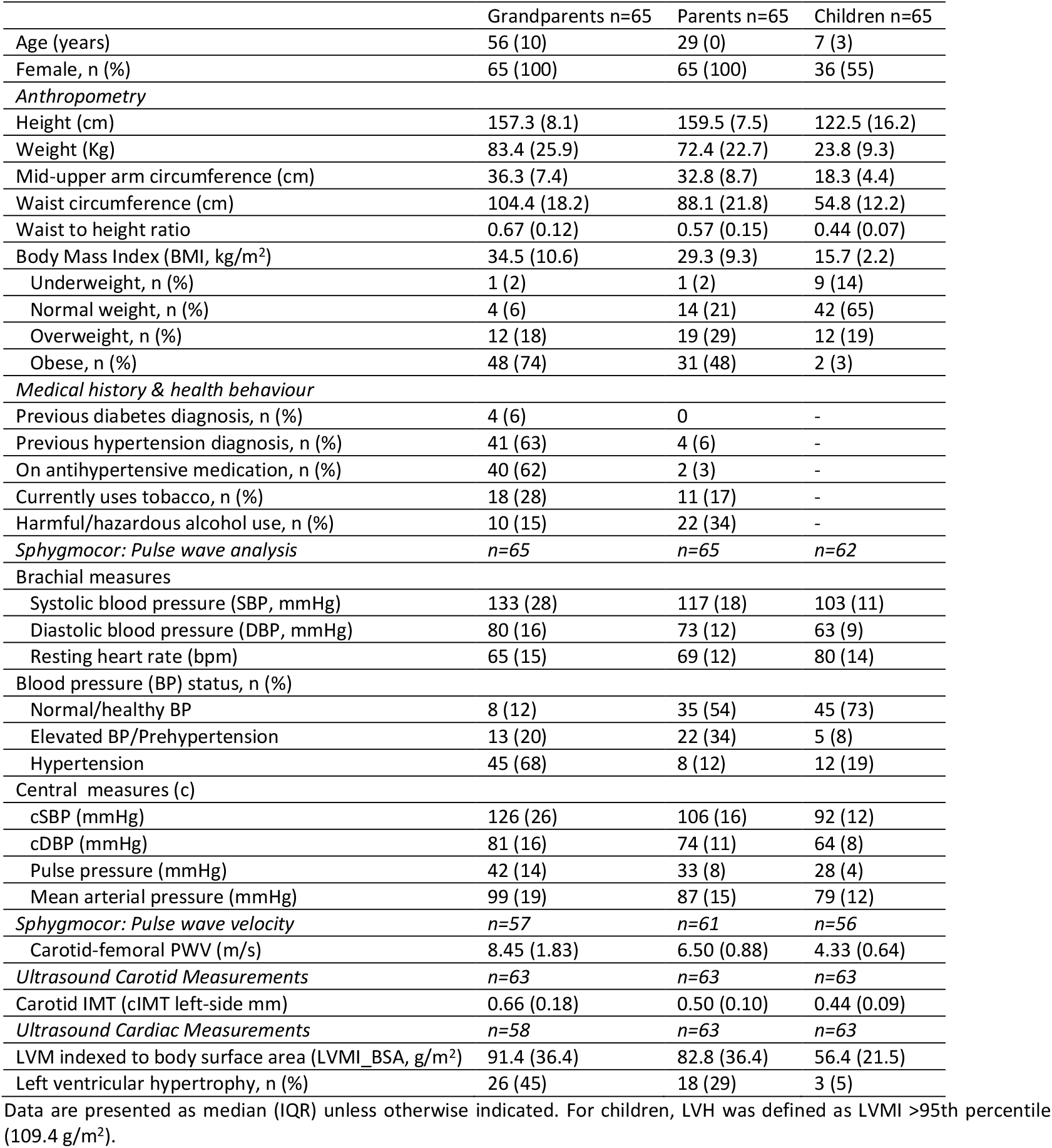
Characteristics of the n=65 included families (grandparents, parents and children)

Median age of grandparents, parents and children was 56 years, 29 years and 7 years respectively. Among adults, 92% of grandparents and 77% of parents were overweight or obese. While the majority of children were a healthy weight (65%), one in five was overweight or obese. Elevated BP (pre-hypertension or hypertension) was present in 88% of grandparents, 46% of parents, and 27% of children. In general, markers of cardiovascular disease risk worsened with age (**Table 1**), with 5% of children, 29% of parents, and 45% of grandparents categorised as having left ventricular hypertrophy.

### Results of random family and heritability analysis

**Table 2** shows the results from comparing biological family pairs to randomly generated non-biological pairings, with statistically significant associations observed within families for cIMT between grandparents and parents, and for LVMI between all first-degree generations. Combining the heritability estimates from the different methods (**Table 3**) showed that heritability of cIMT ranged from 0.234 to 0.439 such that between 23% and 44% of the variation in cIMT was explained by heritability within families. For LVMI, the estimates from the various methods were closer, suggesting between 21% and 39% of the variation in LVMI was explained by heritability within families. Importantly, though the heritability estimates from the different estimation methods were related (**Suppl. figure 1**) and each parameter overlapped, high standard deviation for phylogenetic variance estimates as well as heritability estimates were observed.

**Table 2.** Results of random family analysis.

| Outcome | Exposure | Observed effect* [T(obs)] | c | n | P =c/n |
| --- | --- | --- | --- | --- | --- |
| Brachial SBP - GC | Brachial SBP - GP | 0.029 | 3123 | 5000 | 0.625 |
| Brachial SBP - GC | Brachial SBP - P | 0.123 | 1027 | 5000 | 0.205 |
| Brachial SBP - P | Brachial SBP - GP | 0.109 | 967 | 5000 | 0.193 |
| Brachial DBP - GC | Brachial DBP - GP | -0.006 | 4647 | 5000 | 0.929 |
| Brachial DBP - GC | Brachial DBP - P | 0.063 | 2676 | 5000 | 0.535 |
| Brachial DBP - P | Brachial DBP - GP | 0.001 | 4970 | 5000 | 0.994 |
| Central SBP - GC | Central SBP - GP | -0.005 | 4649 | 5000 | 0.930 |
| Central SBP - GC | Central SBP - P | 0.075 | 2249 | 5000 | 0.450 |
| Central SBP - P | Central SBP - GP | 0.094 | 1392 | 5000 | 0.278 |
| Central DBP - GC | Central DBP - GP | 0.028 | 3379 | 5000 | 0.676 |
| Central DBP - GC | Central DBP - P | 0.119 | 1180 | 5000 | 0.236 |
| Central DBP - P | Central DBP - GP | 0.006 | 4702 | 5000 | 0.940 |
| PWV - GC | PWV - GP | -0.006 | 4655 | 5000 | 0.931 |
| PWV - GC | PWV - P | 0.166 | 766 | 5000 | 0.153 |
| PWV - P | PWV - GP | 0.104 | 1038 | 5000 | 0.208 |
| cIMT - GC | cIMT - GP | 0.093 | 962 | 5000 | 0.192 |
| cIMT - GC | cIMT - P | 0.171 | 1445 | 5000 | 0.289 |
| cIMT - P | cIMT - GP | 0.133 | 83 | 5000 | **0.017** |
| LVMI_BSA - GC | LVMI_BSA - GP | -0.076 | 2301 | 5000 | 0.460 |
| LVMI_BSA - GC | LVMI_BSA - P | 0.242 | 213 | 5000 | **0.043** |
| LVMI_BSA - P | LVMI_BSA - GP | 0.277 | 102 | 5000 | **0.020** |

GC- grandchild, P- parent, GP- grandparent, SBP – systolic blood pressure, DBP – diastolic blood pressure, cIMT – carotid intima media thickness, LVMI\_BSA – left ventricular mass indexed to body surface area. \*All exposure effects were adjusted for age, height, weight and sex in the regression models. P: the empirical probability value. C: the number of absolute effects ≥ the observed targeted generation effect (e.g. grandparent on grandchild, grandparent on parent etc. as indicated by the formula below). n: the number of generated pseudo random families assessed on the targeted generation effect to determine c, where: $c = \#\{ |T| \geq |T(\text{obs})| \}$

**Table 3.** Heritability estimates from different methods

|  | cIMT (mm) |  |  | LVMI_BSA (g/m<sup>2</sup>) |  |  |
| --- | --- | --- | --- | --- | --- | --- |
|  | ReML<sup>1</sup> | MCMC<sup>2</sup> | HMC<sup>3</sup> | ReML | MCMC | HMC |
| Phylogenetic variance (p) | 0.131 (0.114) | 0.310 (0.101) | 0.175 (0.111) | 0.180 (0.172) | 0.405 (0.141) | 0.240 (0.154) |
| Error variance | 0.426 (0.070) | 0.385 (0.056) | 0.416 (0.065) | 0.660 (0.107) | 0.603 (0.085) | 0.647 (0.095) |
| Phenotypic variance | 0.556 (0.080) | 0.695 (-) | 0.591 (-) | 0.840 (0.122) | 1.008 (-) | 0.887 (-) |
| Heritability ($h^2$) | 0.234 (0.179) | 0.439 (0.098) | 0.282 (0.146) | 0.214 (0.182) | 0.394 (0.099) | 0.258 (0.139) |
| $\beta^4$ | 0.709 (0.048) | 0.705 (0.048) | 0.708 (0.047) | 0.496 (0.059) | 0.496 (0.059) | 0.493 (0.060) |

<sup>1</sup>Restricted Maximum Likelihood, <sup>2</sup>Markov Chain Monte Carlo method, <sup>3</sup>Hamiltonian Monte Carlo method, <sup>4</sup>coefficient for age which was adjusted for in all models for both vascular markers

## Discussion

The aim of this study was to examine a range of phenotypic markers of cardiovascular risk across three generations to determine the degree to which these measures of vascular health are transmitted through generations in an urban South African family cohort, and give an indication of whether these findings in older generations can be used to trigger assessments of cardiovascular risk in younger generations. While we did not find significant heritability of blood pressure, possibly due to the high prevalence of elevated blood pressure and hypertension across all generations, our results do suggest that, in this population, structural markers of CV risk (intima media thickness in the common carotid artery (cIMT) and left ventricular mass (LVMI)) are heritable across African generations. This supports the intergenerational transmission of cardiovascular risk and identifies potential markers for the detection of at risk families.

To our knowledge, there is scant information to date on the degree to which these phenotypic markers of cardiovascular risk are heritable within African families. However, the heritability estimates we identified for these structural cardiovascular markers are similar to those reported in several previous studies from research outside of Africa. For example, our estimates for heritability of cIMT (23-44%) are similar to the 38% heritability reported in 586 families from the Framingham heart study^19^ and the 34% reported in Latino parent-offspring pairs (69 families)^50^. However, our estimates are lower than the 56% heritability reported from 100 Dominican families in the Northern Manhattan study^51^ and slightly higher than the 21% estimate reported in 32 American Indian families from the Strong Heart Family study^17^. Lower estimates may be related to the pedigrees included in the samples. For example, the Strong Heart Family study included first, second, third, fourth and greater degree relatives; while the other studies included only first degree relatives. Further studies in first-degree relatives from 76 families in France provide a similar cIMT heritability estimate of 30%^52^. Given our finding that significant heritability was observed in first-degree relatives (grandparent-parent) our results broadly agree with other studies and may be among the first to identify this heritability in families in Africa.

We also saw broad agreement between our heritability estimates for LVMI (21% to 39%) with estimates from studies outside of Africa including the Framingham heart study (30% heritability between parent-child pairs)^22^, from 52 White European families (23%), and from 368 Chinese families living in Taiwan (27%)^21 53^. Again, our estimate is higher than that from the Strong Heart Study (17%)^54^ and lower than that from the Northern Manhattan study (49%)^55^. Our estimates are also lower than those from 169 hypertensive Japanese families living in Hawaii (43%)^56^ and from the HyperGEN study (46%; 527 families, 51% African-America; 53% hypertensive)^57^. Generally, these higher heritability estimates for LVMI are from studies including or exclusively involving hypertensive participants. However, this may not in itself explain the higher estimates as we included family members with hypertension, as did the GENOA study in African-American hypertensive siblings with 34% estimated heritability of LVMI^24^, falling within the range of our findings.

When comparing our LVMI heritability estimates with the one study found within Africa (from 181 nuclear families in our same urban township in South Africa)^26^, our estimates are lower. However, this study indexed LVM to height rather than BSA, with other studies showing this produces higher indexed LVMI values^58^. Importantly, the agreement between the studies that LVMI is heritable within families in this region supports the need for improved screening services.

Our findings for blood pressure were not expected and are contrary to other studies where blood pressure heritability has been observed within families. In a systematic review and meta-analysis by Kolifarhood et al. (2019), heritability of SBP and DBP was observed across regions ranging from 17-52% for SBP and 19-41% for DBP, though estimates were lower in African populations^59^. However, African data were scarce with one study in Nigeria from Adeyemo et al. (2002) reporting heritability estimates of 34% for SBP and 29% for DBP in 528 families including 1825 individuals^60^. While this was a large sample, heritability of BP has been observed in smaller African studies. For example, Bochud et al. (2005) found a significant heritability estimate for office SBP of 28% in 314 East African (Seychellois) adults from 76 families^30^. However, in this study family members were recruited for having at least two siblings with hypertension and family relationships included first degree (sibling pairs, parent-offspring pairs), second degree (grandparent-grandchild pairs, avuncular pairs i.e. uncle/aunt-niece/nephew) and third degree (first cousin pairs) relatives. Our research included only first and second degree relatives in whom heritability might be expected to be higher, though our overall sample size (n=198) was smaller.

We also expected to find significant heritability for arterial stiffness within our families. Data from the Framingham Heart Study (1480 individuals from 817 families) suggests around 40% heritability of carotid-femoral pulse wave velocity^20^. While evidence from a study in Brazil (125 families, 1675 individuals) shows a lower heritability estimate (27%)^61^, though this study also included first, second and third degree relatives. To our knowledge, our results may be some of the first to investigate the intergenerational heritability of carotid-femoral pulse wave velocity as a measure of arterial stiffness in families within South Africa and possibly, in Africa highlighting the need for further work in African families, perhaps increasing sample size through the inclusion of third-degree relatives.

Given constrained resources for cardiovascular disease treatment in the region, pragmatic and targeted prevention approaches are needed leveraging measurements that may be taken as part of routine clinical practice. Given the heritability of the factors identified in this study, we are not suggesting that people should be screened for these factors to identify at risk children and families. Rather that offspring of adults in whom these factors are found should be targeted for rigorous assessment of risk, especially for raised LVM where this is measured in clinical practice.

### Strengths & Limitations

Our findings must be viewed in light of the limitations of this research, most notably the small sample size resulting in high standard deviations observed for phylogenetic variance estimates as well as heritability estimates. However, our heritability estimates from the different estimation methods for each parameter overlap giving confidence for our analysis, and the heritability estimates observed for CIMT and LVMI are similar to many of those reported previously. Additionally, the number of families included in this analysis is similar or more than many other heritability studies, with the random family method increasing the numbers of comparisons available. While our findings contribute to the small but growing evidence base for Africa, further research is needed across the continent to assess the generalisability of our results.

A further limitation results from the individuals in which we could not collect all phenotypic markers of cardiovascular risk, most notably the SphygmoCor PWV and the echocardiography measures. This difficulty was in part due to excess body mass, for example the mean adult BMI of those with unsuccessful echocardiography measurement was 40.9 ± 10.5 kg/m^2^. Our lack of 24 hour ambulatory blood pressure monitoring (ABPM) data within families is also a limitation and future studies should consider the use of ABPM where feasible, as heritability estimates appear higher for ABPM than for office BP^62^. While we have successfully utilised ABPM in South African adults previously^63^, this was significantly more challenging in this urban cohort with young children and our attempts were not successful. Community-based support for families during ABPM measurement may be helpful in the future.

While it is noted that comparison with other studies can be problematic due to different populations, methods, study designs, and environmental influence on phenotypic variance as highlighted by North et al. (2002)^17^, we have taken care to compare our results only to studies that are methodologically similar. For example, all comparisons for LVMI heritability presented here include only studies using echocardiographic measurement of LVM, as LVM heritability estimates from electrocardiography may be higher^23^. Furthermore, heritability estimates for IMT often vary between the common carotid artery (CCA) and the internal carotid (ICA), with heritability estimates frequently higher for CCA, so that it is important to compare results for IMT measured in the same location.

A key strength of this research is the contribution of evidence for the heritability and intergenerational transmission of cardiovascular health in African families, including children prior to adolescence, and the comparison of several different methods to estimate heritability. Further, the high levels of elevated blood pressure and hypertension observed in our population across older and younger adults and in the children reinforce the need for prevention programmes early in life.

## Conclusion

Our results suggest that structural cardiovascular indices in the common carotid artery and in the left ventricle of the heart are heritable within African families. Where adults are identified with elevated carotid intima media thickness or left ventricular hypertrophy, screening should be conducted in first and second-degree relatives, especially to identify younger individuals most at risk of later poor vascular health, where prevention efforts may yield the greatest returns. Better understanding of the factors that promote transmission of poor vascular health from one generation to the next will support development of interventions to break the upward spiral of CVD on the continent.

## Data Availability

All data produced in the present study are available upon reasonable request to the authors

## Contributorship statement

LJW, JD, SAN and IM conceived the idea for the manuscript and designed the analyses. IM and LJW performed the analyses. LJW, IM, JD, AKR, LS, SC, WS, SAN all contributed to the interpretation of the results. All authors contributed to drafting the manuscript and have seen and approved the final version. LJW is the guarantor for this work and accepts full responsibility for the work.

## Data sharing statement

Data is available on request from SAN.

---

**Supplementary figure 1.**
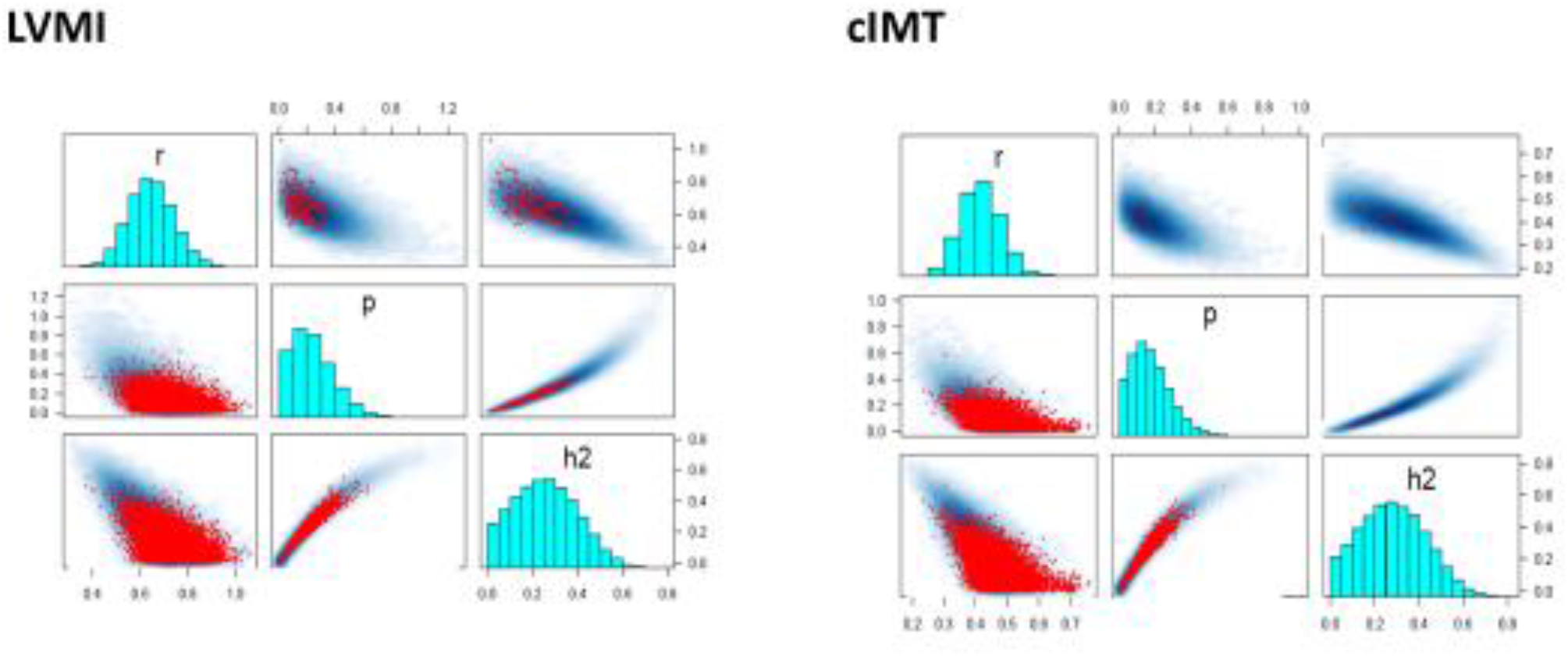
showing the relationships between the heritability parameters for LVMI (adjusted for body surface areas) and carotid IMT (cIMT).

## Appendix

*Additional information on cardiovascular assessment methods*

Arterial stiffness (carotid-femoral pulse wave velocity - PWV) was estimated, with tonometry of the carotid artery during inflation of an appropriate size femoral cuff. Pulse wave analysis (for central pressure estimation) and PWV measurement were set at 10 second intervals. Duplicate measures of PWV were taken and if the difference between PWV measures was ≥0.5 m/s, a third measure was taken and the average of two readings within 0.5m/s of each other used for analysis. All measures were taken on the right side with the participant resting supine for 10 minutes prior to measurement, and using the direct distance method to estimate aortic path length^1^. A total of 4 trained operators performed the PWV measurements after confirming inter-observer variability was acceptable (< 0.5 m/s).

Left ventricular mass (LVM) was measured in 2D mode with transthoracic echocardiography following the American Society of Echocardiography (ASC) protocol^2^. The 2D mode has been shown to be superior to M-Mode for studies of LVM within families^3^. LV mass was assessed at end-diastole perpendicular to the long axis of the left ventricle. The Devereux formula was used to calculate LVM: LVM (g) = 0.8 × 1.04 ((LVDd + IVSd + LVPWd)3 – LVDd3) + 0.6 where LVDd=left ventricular diastolic diameter; IVSd= intraventricular septal diameter, LVPWd= left ventricular posterior wall thickness in diastole^4^.

Carotid intima-media thickness (cIMT) was determined using high resolution B-mode ultrasound employing a linear array 7.5 MHz probe as recommended^5^. Images of at least 1 cm length were obtained of the far wall of the distal portion of the left common carotid artery (CCA) from an optimal angle of incidence (defined as the longitudinal angle of approach where both branches of the internal and external carotid artery are visualised simultaneously). Semi-automated border detection and quality control software were used to calculate cIMT, with at least 3 measurements obtained from the left side and the mean used for analysis. Previous studies have reported no major differences between left and right CCA IMT in associations with cardiovascular disease^6^. All ultrasound measures were taken with the Mindray DC-70 Ultrasound system (Mindray, Shenzen China).

## References

1. World Health Organization - Noncommunicable Diseases (NCD) Country Profile; South Africa, 2018:https://www.who.int/nmh/countries/zaf_en.pdf?ua=1.

2. United Nations - World Population Prospects: South Africa Life Expectancy 1950-2021:https://www.macrotrends.net/countries/ZAF/south-africa/life-expectancy.

3. National Department of Health, Statistics South Africa, South African Medical Research Council, ICF: South Africa Demographic and Health Survey 2016.

4. Owolabi M, Miranda JJ, Yaria J, et al. Controlling cardiovascular diseases in low and middle income countries by placing proof in pragmatism. BMJ Global Health 2016;1(3):e000105. doi: 10.1136/bmjgh-2016-000105

5. Sailam V, Karalis DG, Agarwal A, et al. Prevalence of emerging cardiovascular risk factors in younger individuals with a family history of premature coronary heart disease and low Framingham risk score. Clinical Cardiology: An International Indexed and Peer-Reviewed Journal for Advances in the Treatment of Cardiovascular Disease 2008;31(11):542–45.

6. Lorenz MW, Markus HS, Bots ML, et al. Prediction of clinical cardiovascular events with carotid intima-media thickness: a systematic review and meta-analysis. Circulation 2007;115(4):459–67.

7. Aznaouridis K, Dhawan SS, Quyyumi AA. Cardiovascular Risk Prediction by Measurement of Arterial Elastic Properties and Wall Thickness. Advances in Vascular Medicine: Springer 2009:399–421.

8. Vlachopoulos C, Aznaouridis K, Stefanadis C. Prediction of cardiovascular events and all-cause mortality with arterial stiffness: a systematic review and meta-analysis. Journal of the American College of Cardiology 2010;55(13):1318–27.

9. Gosse P. Left ventricular hypertrophy as a predictor of cardiovascular risk. Journal of hypertension 2005;23:S27–S33.

10. Peters SA, Huxley RR, Woodward M. Comparison of the sex-specific associations between systolic blood pressure and the risk of cardiovascular disease: a systematic review and meta-analysis of 124 cohort studies, including 1.2 million individuals. Stroke 2013;44(9):2394–401.

11. Kagura J, Ong KK, Adair LS, et al. Paediatric hypertension in South Africa: An underestimated problem calling for action. SAMJ: South African Medical Journal 2018;108(9):708–09.

12. Hottenga J-J, Boomsma DI, Kupper N, et al. Heritability and stability of resting blood pressure. Twin Research and Human Genetics 2005;8(5):499–508.

13. Snieder H, Harshfield GA, Treiber FA. Heritability of blood pressure and hemodynamics in African-and European-American youth. Hypertension 2003;41(6):1196–201.

14. Kolifarhood G, Daneshpour M, Hadaegh F, et al. Heritability of blood pressure traits in diverse populations: a systematic review and meta-analysis. Journal of human hypertension 2019;33(11):775–85.

15. Djami-Tchatchou AT, Norton GR, Redelinghuys M, et al. Intrafamilial aggregation and heritability of office− day blood pressure difference in a community of African ancestry: implications for genetic association studies. Blood pressure monitoring 2014;19(6):346–52.

16. Tarnoki AD, Tarnoki DL, Stazi MA, et al. Heritability of central blood pressure and arterial stiffness: a twin study. J Hypertens 2012;30(8):1564–71. doi: 10.1097/HJH.0b013e32835527ae [Published online First: 2012/06/13]

17. North KE, MacCluer JW, Devereux RB, et al. Heritability of carotid artery structure and function: the Strong Heart Family Study. Arteriosclerosis, thrombosis, and vascular biology 2002;22(10):1698–703.

18. Moskau S, Golla A, Grothe C, et al. Heritability of carotid artery atherosclerotic lesions: an ultrasound study in 154 families. Stroke 2005;36(1):5–8.

19. Fox CS, Polak JF, Chazaro I, et al. Genetic and environmental contributions to atherosclerosis phenotypes in men and women: heritability of carotid intima-media thickness in the Framingham Heart Study. Stroke 2003;34(2):397–401.

20. Mitchell GF, DeStefano AL, Larson MG, et al. Heritability and a genome-wide linkage scan for arterial stiffness, wave reflection, and mean arterial pressure: the Framingham Heart Study. Circulation 2005;112(2):194–99.

21. Jin Y, Kuznetsova T, Bochud M, et al. Heritability of left ventricular structure and function in Caucasian families. European journal of echocardiography 2011;12(4):326–32.

22. Post WS, Larson MG, Myers RH, et al. Heritability of left ventricular mass: the Framingham Heart Study. Hypertension 1997;30(5):1025–28.

23. Mayosi B, Keavney B, Kardos A, et al. Electrocardiographic measures of left ventricular hypertrophy show greater heritability than echocardiographic left ventricular mass. European heart journal 2002;23(24):1963–71.

24. Fox ER, Klos KL, Penman AD, et al. Heritability and genetic linkage of left ventricular mass, systolic and diastolic function in hypertensive African Americans (From the GENOA study). American journal of hypertension 2010;23(8):870–75.

25. Laurent S, Parati G. Heritability of arterial stiffness and central blood pressure: the Holy Grail for detecting patients at high cardiovascular risk? Journal of Hypertension 2012;30(8)

26. Peterson VR, Norton GR, Redelinghuys M, et al. Intrafamilial aggregation and heritability of left ventricular geometric remodeling is independent of cardiac mass in families of African ancestry. American journal of hypertension 2015;28(5):657–63.

27. Peterson VR, Norton GR, Libhaber CD, et al. Intrafamilial aggregation and heritability of tissue Doppler indexes of left ventricular diastolic function in a group of African descent. Journal of the American Society of Hypertension 2016;10(6):517-26. e11.

28. Richter LM, Norris SA, De Wet T. Transition from Birth to Ten to Birth to Twenty: the South African cohort reaches 13 years of age. Paediatric and perinatal epidemiology 2004;18(4):290–301.

29. Richter LM, Naicker SN, Norris SA. Birth to Twenty Plus: Early health and development from a longitudinal perspective. Child and Adolescent Development: An expanded focus on public health in Africa 2018:114.

30. Bochud M, Bovet P, Elston RC, et al. High heritability of ambulatory blood pressure in families of East African descent. Hypertension 2005;45(3):445–50.

31. Klein TW. Heritability and genetic correlation: statistical power, population comparisons, and sample size. Behavior Genetics 1974;4(2):171–89.

32. Von Elm E, Altman DG, Egger M, et al. The Strengthening the Reporting of Observational Studies in Epidemiology (STROBE) statement: guidelines for reporting observational studies. Bulletin of the World Health Organization 2007;85:867–72.

33. Global Adult Tobacco Survey Collaborative Group. Tobacco Questions for Surveys: A Subset of Key Questions from the Global Adult Tobacco Survey (GATS), 2nd Edition. Atlanta, GA: Centers for Disease Control and Prevention, 2011.

34. Babor TF, Higgins-Biddle JC, Saunders JB, Monteiro MG; The Alcohol Use Disorders Identification Test, Guidelines for Use in Primary Care, Second Edition, Department of Mental Health and Substance Dependence, World Health Organization, CH-1211 Geneva 27, Switzerland. ©World Health Organization 2001.

35. World Health Organization. Waist circumference and waist-hip ratio: report of a WHO expert consultation, Geneva, 8-11 December 2008. 2011

36. World Health Organization Physical status: The use of and interpretation of anthropometry, Report of a WHO Expert Committee: World Health Organization 1995.

37. Cole TJ, Bellizzi MC, Flegal KM, et al. Establishing a standard definition for child overweight and obesity worldwide: international survey. Bmj 2000;320(7244):1240.

38. Obe MA. Waist to height ratio and the Ashwell® shape chart could predict the health risks of obesity in adults and children in all ethnic groups. Nutrition & Food Science 2005

39. Flynn JT, Kaelber DC, Baker-Smith CM, et al. Clinical practice guideline for screening and management of high blood pressure in children and adolescents. Pediatrics 2017;140(140)

40. Devereux RB, Alonso DR, Lutas EM, et al. Echocardiographic assessment of left ventricular hypertrophy: comparison to necropsy findings. The American journal of cardiology 1986;57(6):450–58.

41. Mosteller R. Simplified calculation of body-surface area. The New England journal of medicine 1987;317(17):1098–98.

42. Shapiro SS, Wilk MB. An analysis of variance test for normality (complete samples). Biometrika 1965;52(3/4):591–611.

43. Heß S. Randomization inference with Stata: A guide and software. The Stata Journal 2017;17(3):630–51.

44. Usuzaki T, Chiba MSS, Hotta S. Random family method: Confirming inter-generational relations by restricted re-sampling. arXiv preprint arXiv:200203467 2020

45. Hadfield JD, Wilson AJ, Garant D, et al. The misuse of BLUP in ecology and evolution. The American Naturalist 2010;175(1):116–25.

46. Morrissey M, Kruuk L, Wilson AJ. The danger of applying the breeder’s equation in observational studies of natural populations. Journal of evolutionary biology 2010;23(11):2277–88.

47. Zhao JH, Congdon P. Bayesian linear mixed models with polygenic effects. Journal of Statistical Software 2018;85(1):1–27.

48. Zhao J, Zhao H, Zhu L. Pivotal variable detection of the covariance matrix and its application to high-dimensional factor models. Statistics and Computing 2018;28(4):775–93.

49. Sinnwell JP, Therneau TM, Schaid DJ. The kinship2 R package for pedigree data. Human heredity 2014;78(2):91–93.

50. Xiang AH, Azen SP, Buchanan TA, et al. Heritability of Subclinical Atherosclerosis in Latino Families Ascertained Through a Hypertensive Parent. Arteriosclerosis, Thrombosis, and Vascular Biology 2002;22(5):843–48. doi: doi:10.1161/01.ATV.0000015329.15481.E8

51. Sacco RL, Blanton SH, Slifer S, et al. Heritability and linkage analysis for carotid intima-media thickness: the family study of stroke risk and carotid atherosclerosis. Stroke 2009;40(7):2307–12.

52. Zannad F, Visvikis S, Gueguen R, et al. Genetics strongly determines the wall thickness of the left and right carotid arteries. Human genetics 1998;103(2):183–88.

53. Chien K-L, Hsu H-C, Su T-C, et al. Heritability and major gene effects on left ventricular mass in the Chinese population: a family study. BMC cardiovascular disorders 2006;6(1):1–9.

54. Bella JN, MacCluer JW, Roman MJ, et al. Heritability of left ventricular dimensions and mass in American Indians: The Strong Heart Study. Journal of hypertension 2004;22(2):281–86.

55. Hank Juo S-H, Di Tullio MR, Lin H-F, et al. Heritability of left ventricular mass and other morphologic variables in Caribbean Hispanic subjects: the Northern Manhattan Family Study. Journal of the American College of Cardiology 2005;46(4):735–37.

56. Assimes TL, Narasimhan B, Seto TB, et al. Heritability of left ventricular mass in Japanese families living in Hawaii: the SAPPHIRe Study. Journal of hypertension 2007;25(5):985–92.

57. De Simone G, Tang W, Devereux RB, et al. Assessment of the interaction of heritability of volume load and left ventricular mass: the HyperGEN offspring study. Journal of hypertension 2007;25(7):1397–402.

58. Hense H-W, Gneiting B, Muscholl M, et al. The associations of body size and body composition with left ventricular mass: impacts for indexation in adults. Journal of the American College of Cardiology 1998;32(2):451–57. doi: doi:10.1016/S0735-1097(98)00240-X

59. Kolifarhood G, Daneshpour M, Hadaegh F, et al. Heritability of blood pressure traits in diverse populations: a systematic review and meta-analysis. Journal of Human Hypertension 2019;33(11):775–85. doi: 10.1038/s41371-019-0253-4

60. Adeyemo AA, Omotade OO, Rotimi CN, et al. Heritability of blood pressure in Nigerian families. Journal of Hypertension 2002;20(5):859–63.

61. Alvim RO, Horimoto AR, Oliveira CM, et al. Heritability of arterial stiffness in a Brazilian population: Baependi Heart Study. Journal of hypertension 2017;35(1):105–10.

62. Fava C, Burri P, Almgren P, et al. Heritability of ambulatory and office blood pressure phenotypes in Swedish families. Journal of hypertension 2004;22(9):1717–21.

63. Ware LJ, Rennie KL, Gafane LF, et al. Masked hypertension in low-income South African adults. The Journal of Clinical Hypertension 2016;18(5):396–404.

## References

1. Van Bortel LM, Laurent S, Boutouyrie P, et al. Expert consensus document on the measurement of aortic stiffness in daily practice using carotid-femoral pulse wave velocity. Journal of hypertension 2012;30(3):445–48.

2. Lang RM, Badano LP, Mor-Avi V, et al. Recommendations for cardiac chamber quantification by echocardiography in adults: an update from the American Society of Echocardiography and the European Association of Cardiovascular Imaging. European Heart Journal-Cardiovascular Imaging 2015;16(3):233–71.

3. Shah SJ, Ober C, Lang RM. Two-Dimensional Echocardiography Is Superior to M-mode for the Determination of Left Ventricular Mass: Evidence from a Heritability Study. Journal of Cardiac Failure 2007;13(6):S155.

4. Devereux RB, Alonso DR, Lutas EM, et al. Echocardiographic assessment of left ventricular hypertrophy: comparison to necropsy findings. The American journal of cardiology 1986;57(6):450–58.

5. Touboul P-J, Hennerici M, Meairs S, et al. Mannheim carotid intima-media thickness and plaque consensus (2004–2006–2011). Cerebrovascular diseases 2012;34(4):290–96.

6. Bots ML, de Jong PT, Hofman A, et al. Left, right, near or far wall common carotid intima-media thickness measurements: associations with cardiovascular disease and lower extremity arterial atherosclerosis. Journal of clinical epidemiology 1997;50(7):801–07.

